# What adults in rural South Asia eat and when they eat it: evidence from Bangladesh, India, and Nepal

**DOI:** 10.1101/2025.01.06.25320064

**Authors:** Samuel Scott, Sharvari Patwardhan, Marie Ruel, Suman Chakrabarti, Sumanta Neupane, Swetha Manohar, Mourad Moursi, Purnima Menon

**Affiliations:** International Food Policy Research Institute; FHI Solutions

## Abstract

**Background:** Poor diets pose a threat to all forms of malnutrition and diet-related noncommunicable diseases (NCDs). Data on dietary patterns are scarce in South Asia.

**Objectives:** We sought to describe overall diet quality, intake of foods and food groups, and eating occasions among adults in rural South Asia.

**Methods:** Data were from five districts across Bangladesh (n=2,802 individuals), India (n=1,672), and Nepal (n=1,451). The Global Diet Quality Score (GDQS) application was used to measure intake of foods on the previous day, with each food tagged to an eating occasion. Diet quality and the risk of diet-related noncommunicable diseases (NCDs) were described using GDQS total (0–49), GDQS positive (0–32) and GDQS negative (0–17) metrics for overall, healthy, and unhealthy food intake respectively.

**Results:** Diet quality was low, with similar scores across countries for GDQS total (17-19 depending on country), GDQS positive (7–8) and GDQS negative (10–12), indicating low intake of healthy foods as the main contributor to poor diets. Over 90% of adults had levels of GQDS scores associated with moderate/high risk for diet-related NCDs, with the proportion at high risk in Bangladesh being 2-3x higher than other countries. Across sites, intake of refined grains (white rice), sweets (sugar, biscuits), and white tubers (potatoes) was common. One-third of adults did not eat breakfast in Nepal, and snacking was twice as common in males (63%) versus females (33%) in Bangladesh.

**Conclusions:** These findings highlight the need to improve diets in rural South Asia and may help inform interventions targeting food intake patterns.

## INTRODUCTION

Unhealthy diets underpin all forms of malnutrition and were estimated to be responsible for 11 million deaths and 255 million disability-adjusted life years in 2017 (1). South Asia, home to a quarter of the global population and over 40% of the population living in poverty, is one of two global regions (the other being Sub-Saharan Africa) where diets did not improve from 1990 to 2018 (2). Unhealthy diets also lead to productivity losses (3–5) and increased greenhouse gas emissions (6), thus have economic and planetary consequences that extend beyond human health. The constituency of a healthy diet varies by context and individual, but they include a diversity of foods – fruits, vegetables, legumes, nuts, whole grains and varying amounts of animal-source foods – and limit consumption of foods high in sugar, salt and fat (7,8).

There is a dearth of dietary data available from low-and-middle income countries and from South Asia in particular (9,10). Understanding what people eat and when they eat throughout the day can uncover potential entry points for targeted dietary and food systems interventions. In a recent review of evidence on behavior change interventions to address unhealthy food intake, only 5 of 145 (or 3% of) studies were from South Asia (11), underscoring the limited evidence base to inform the design of such interventions in the region.

Measurement challenges are a major barrier to collection of dietary data (12). Most population-based surveys that measure food intake rely on measurement of the types and amounts of foods purchased and consumed by a household over a given period of time - or household expenditure on food. While these tools can be useful for assessing the foods available and consumed by the household, they do not capture individual-level food intake or the timing of intake over 24 hours. While numerous studies have used the 24-hour recall approach to measure individual diet -- where the respondent provides details on the type and quantity of each food/ ingredient consumed, over one or multiple days, this method is time-consuming and expensive and carries a high respondent burden. To overcome some of these challenges, The Global Diet Quality Score (GDQS) application was developed to operationalize the collection of a diet quality metric, the GDQS. This method captures intake of all foods/ingredients in the past 24 hours, with quantity estimated at the food group rather than ingredient level, which reduces the interview length compared to traditional 24-hour recall. Importantly, GDQS has been validated for adults using data from 15 countries, including India, and results showed that the GDQS performed as well as the minimum Dietary Diversity – Women indicator in predicting nutrient intake and indicators of undernutrition; the GDQS also performed better than the Alternative Health Eating Index – 2010 in terms of predicting metabolic syndrome, change in waist circumference, and incident type 2 diabetes (13,14), making this tool a solid alternative to the 24-h recall for measuring the quality of diets at the population level.

Following the High Level Panel of Experts food systems framework (15), which places consumer diets as one of the main outcomes of food systems processes, we conducted a holistic assessment of district-level agrifood systems in five districts in rural Bangladesh, India, and Nepal. Under the Consultative Group for International Agricultural Research (CGIAR) initiative ‘Transforming Agrifood Systems in South Asia’ (TAFSSA), these assessments took a plate-to-farm approach, including measurement on diets, markets, food production, climate, and gender, among other factors. The aims of the current paper are to describe 1) what foods adult males and females in rural South Asia are eating and 2) patterns and timing of food intake by gender. The dearth of information on food intake patterns for this rural population prevents policy action.

## METHODS

### Sites, sampling, and respondents

Data were collected in five locations in February-May 2023 and included Nalanda district in Bihar state (India); Surkhet district in Lumbini Province and Banke district in Karnali Province (Nepal); and Rangpur district in the Rangpur division and Rajshahi district in the Rajshahi Division (Bangladesh). These locations were selected because they are recognized hot spots for challenges related to poverty, climate, malnutrition and social inclusion. The samples are representative of all rural households in a district with an adolescent. The sampling approach and sample size determination followed the example of India’s equivalent of the Demographic and Health Surveys, the National Family Household Survey (NFHS), a population-based survey representative at the district level most recently conducted in 2019-2021. We erred on the higher end of the number of households per district in NFHS and used a sample size of 1000 households per district for our survey. In Nepal, relative to Bangladesh and India, districts were much less populated, thus we included 500 households per district in Nepal. These sample sizes exceed recommendations for the number of respondents required to guarantee precision in quantitative 24-hour dietary recall surveys (16).

The primary sampling unit (PSU) was villages (in Bangladesh and India) and wards (in Nepal). PSUs were selected from the most recent National Census datasets from each district within each country, using a Probability Proportional to Size (PPS) sampling strategy. In the first stage of sampling, rural villages or wards were selected at random, with a probability proportional to the number of households that reside in the PSU. In the second stage, a household listing exercise was done and 20 households with adolescent members invited to participate in the study. The rationale for only including households with adolescents was to better understand adolescents’ roles in agrifood systems. One adult female aged 20+ years and one adult male aged 20+ years per household were identified as respondents. The adult female was the female household member primarily responsible for managing the household and the adult male was the male household member primarily responsible for agricultural activities.

Three survey firms supported data collection: Data Analysis and Technical Assistance in Bangladesh, Kabil Professional Services in India, and Institute for Integrated Development studies in Nepal. After explaining the survey’s objectives and procedures, enumerators obtained verbal informed consent from respondents. Participation in the survey was voluntary and participants were free to end interviews at any time by informing survey staff.

The protocol, informed consent forms, and study questionnaires were approved by the Institutional Review Boards (IRB) of the International Food Policy Research Institute in Washington, DC, USA; the Institute of Health Economics, University of Dhaka, Bangladesh; Centre for Media Studies, Delhi, India; and the National Health Research Council, Kathmandu, Nepal.

### GDQS data collection procedure

The GDQS application (17) was pilot tested and the GDQS food database was adapted to each survey context. The Intake – Center for Dietary Assessment / FHI 360 team who developed the application shared a list of 4500 food items based on previous surveys. This list was expanded using existing dietary data from an IFPRI survey in Uttar Pradesh, a state that neighbors Bihar in India, the 2018-2019 Bangladesh Integrated Household Survey, and an inventory of food items prepared for a 24-hour dietary recall survey in Nepal as part of the USAID Suaahara project. After expanding the list to 6000 food items based on existing surveys and pilot testing, the applications instructions and interface were translated to Bangla, Hindi, and Nepali. Enumerator training was conducted separately in each country using the GDQS application and a set of 10 plastic cubes (described below). The GDQS application was then pre-tested and any further context-specific adjustments were incorporated. The GDQS interview start time and end time (in hours, minutes, and seconds) were recorded along with the date of interview. The GDQS interview duration was calculated in minutes as the difference between the end time and the start time.

Seven steps were used to collect data on tablets using the GDQS application. Step 1 involved collecting demographic information about the respondent. In step 2, respondents were asked about everything they ate or drank at home and outside the home on the previous day, from the time they woke up until the time they went to bed and didn’t eat or drink anything more. This information was recorded as foods/dishes/beverages consumed during the following pre-determined list of seven eating occasions: pre-breakfast, breakfast, mid-morning, lunch, afternoon, dinner, and post-dinner. Eating occasions were worded and explained in a way that was understandable in each local context. For each eating occasion, all foods and drinks reported by the respondents were recorded. In case a respondent consumed a mixed dish (a dish prepared using multiple ingredients), they were requested to list the main ingredients of each mixed dish in step 3. Information about spices used in mixed dishes was not collected because spices do not get categorized as a GDQS food group and the quantity of spices used is often unknown or difficult to estimate. In step 4, additional information on certain foods was collected to help classify these ingredients into the GDQS food groups. For instance, if a respondent mentioned that they ate bread, there was a follow-up question asking whether it was white or dark brown to classify bread either in the refined or the whole grains GDQS food group. Step 5 checked whether any of the reported foods were purchased deep fried. Step 6 recorded any sugar or caloric sweeteners consumed that might have been missed in the open recall in step 2. In step 7, respondents were requested to visualize the total amount of foods and beverages consumed belonging to the same food group. A set of ten 3D-printed numbered plastic cubes in a range of predetermined sizes was provided to aid the visualization process. For each food group, the enumerator read back to the respondent all foods belonging to the same food group as classified by the application (based on the open recall in step 2) and asked them to point to the cube size that came closest to the amount of all foods combined belonging to the same food group. For example, if they consumed an apple and a banana (both in the ‘other fruit’ food group) yesterday, they were asked to select the cube number corresponding to the amount of apple and banana combined total volume of the apple plus the banana. Cube sizes were used as a measure of consumed quantity being below, equal to, or above food group–specific cutoffs established in grams, as described in Moursi et al. 2021 (17).

### Dietary metrics

The GDQS is a population-based metric of diet quality which has been validated to predict nutrient inadequacy and non-communicable disease (NCD) risk outcomes globally. The metric consists of 25 food groups: 16 healthy food groups, 2 food groups that are unhealthy when consumed in excess, and 7 unhealthy food groups (**Box 1**) (18). For 24 of the food groups, quantity consumed is classified into: low, medium, and high. For high-fat dairy, quantity of intake is classified into four categories: low, medium, high, and very high. For each respondent, points are assigned based on the levels of intake. For the 16 healthy food groups, more points are given for higher intake.

#### Box 1.

Global Diet Quality Score food groups

Healthy

- Citrus fruits
- Deep orange fruits
- Other fruits
- Dark green leafy vegetables
- Cruciferous vegetables
- Deep orange vegetables
- Other vegetables
- Legumes
- Deep orange tubers
- Nuts and seeds
- Whole grains
- Liquid oils
- Fish and shellfish
- Poultry and game meat
- Low-fat dairy
- Eggs

Unhealthy in excessive amounts

- High-fat dairy
- Red meat

Unhealthy

- Processed meat
- Refined grains and baked goods
- Sweets and ice cream
- Sugar-sweetened beverages
- Juice
- White roots and tubers
- Purchased deep fried foods

For the 7 unhealthy food groups, more points are given for lower intake. For the 2 food groups that are healthy when consumed in moderation, points are given as follows: for the red meat food group, the most points are given for moderate intake (but not low or high intake); for the high fat dairy food group, points are given for moderate and high intake (but not low or very high intake). The list of foods and rankings is provided in **Supplementary Table 1**. The GDQS total metric is a sum of the points across all 25 GDQS food groups, with a possible range of 0 to 49 and a higher score indicating a healthier diet. The GDQS+ sub metric is tabulated by summing points across the 16 healthy GDQS food groups (possible range of 0 to 32). The GDQS– sub metric is tabulated by summing points across the 7 unhealthy GDQS food groups and the 2 GDQS food groups that are unhealthy when consumed in excessive amounts (possible range of 0 to 17). Since more points are awarded for lower intake of unhealthy foods, the GDQS-rewards avoidance of these foods. The total GDQS score is used to classify respondents based on their risk of nutrient inadequacy and diet-related noncommunicable diseases being low (GDQS total ≥23), moderate (≥15 and <23), or high (<15).

### Sociodemographic and socioeconomic data

In addition to dietary data, we collected information on the living conditions and activities of households. Given the focus of the current paper on describing diets, we report on selected factors to provide context to the findings on food intake patterns. Individual level indicators included respondent age and education (years of schooling). Household level indicators included number of members, household head gender, land ownership by females, food insecurity (measured using the food insecurity experiences scale with 8 items and a 30-day recall, using a cutoff of ≥1 to indicate food insecure) (19), water insecurity (water insecurity experiences scale with 4 items and 30-day recall, using a cutoff of ≥4 to indicate water insecure) (20), use of improved drinking water (if the main source of drinking water for household members was piped into dwelling, piped into yard/plot, piped from neighbor, public taps/standpipe, tube well/borehole, protected well, protected spring, rainwater, tanker truck, cart with small tank, bottled water, or community RO plant) and clean cooking fuel (if the type of fuel used by a household was electricity, LPG/natural gas, or biogas), availability of improved toilet facilities (if household members used flush/pour flush toilet, pit latrine, or twin pit/composting toilet), participation in agricultural activities (crop cultivation, livestock and fish rearing), having outstanding loans, and benefiting from social safety net programs (a list of income and food support programs, which varied by country). A household wealth index was generated on the pooled asset ownership data across all districts to allow for comparability of relative wealth. The range of data collected beyond what is reported here, using the plate-to-farm approach, can be found elsewhere (21).

### Statistical analysis

Results are presented in the following order. After describing respondent and household characteristics (mean and SD or %), we first report on indicators of diet quality. These include the mean GDQS scores (GDQS total, GDQS+, GDQS-, and their respective gaps, e.g. since the GDQS+ score is out of 32, a GDQS+ score of 8 will have a GDQS+ gap of 24), the percent of individuals in each GDQS risk category, and two additional indicators from the Global Diet Quality Project (22): the percent of individuals who did not consume any vegetables or fruit on the previous day/night, and the percent who consumed all five food groups typically recommended for daily intake in food based dietary guidelines around the world (at least one fruit, one vegetable, one pulse/nut/seed, one animal-source food, and one starchy staple). Second, we use simple bar charts to show the percentage of males and females who consumed food from each of the 25 GDQS food groups. Third, we show the relative percentage of individuals consuming different food items in healthy and unhealthy food groups using sunburst diagrams. Fourth, spider plots are used to visualize intake of food throughout the day, i.e. the percentage of individuals who consumed any food at each of the seven eating occasions. Lastly, we explore what foods are consumed at each eating occasion using Sankey diagrams.

Analyses were initially conducted at the district level but, given similar results between districts in the same country, data from the districts in Bangladesh (Rajshahi and Rangpur) were combined, as were data from the districts in Nepal (Banke and Surkhet). In addition to district names, country names are provided on tables and figures and referred to in the text for ease of presentation and readability, but we note that results are representative of the study districts, not the entire country, e.g. “females in Bangladesh” should be interpreted as females in Rajshahi and Rangpur districts of Bangladesh. Stata version 18 was used for analysis (23).

## RESULTS

### Respondent and household characteristics

Across the five districts, our sample consisted of 3,992 households, from which we interviewed 3,802 females and 2,123 males (**Table 1**).

**Table 1.** Respondent and household characteristics in Bangladesh, India, and Nepal^1^

|  | Rajshahi, Rangpur<br>(Bangladesh) | Nalanda<br>(India) | Banke, Surkhet<br>(Nepal) |
| --- | --- | --- | --- |
| Respondent characteristics |  |  |  |
| Females, N | 1,864 | 956 | 982 |
| Males, N | 938 | 716 | 469 |
| Female age, yr, Mean (SD) | 37.4 (8.4) | 41.9 (11.6) | 36.8 (9.6) |
| Male age, yr, Mean (SD) | 43.6 (10.9) | 41.9 (11.4) | 45.8 (12.6) |
| Female education, yr, Mean (SD) | 5.0 (3.9) | 2.9 (4.4) | 4.5 (4.3) |
| Male education, yr, Mean (SD) | 4.6 (4.6) | 5.9 (4.9) | 5.2 (4.2) |
| Household characteristics |  |  |  |
| Households, N | 1,992 | 1,000 | 1,000 |
| Household size (SD) | 4.2 (0.02) | 5.8 (0.05) | 4.6 (0.04) |
| Female-headed households, % | 8.6 | 21.0 | 39.4 |
| Land owned by females, % | 19.2 | 12.4 | 30.3 |
| Has improved drinking water source, % | 99.9 | 99.6 | 95.6 |
| Has clean cooking fuel source, % | 29.9 | 77.1 | 77.6 |
| Has improved toilet, % | 98.5 | 41.1 | 97.1 |
| Food insecure<sup>2</sup>, % | 63.8 | 71.6 | 54.9 |
| Water insecure, % | 2.5 | 15.7 | 7.9 |
| Involved in crop cultivation, % | 74.5 | 64.3 | 89.9 |
| Involved in livestock rearing, % | 85.7 | 64.2 | 80.6 |
| Involved in fish rearing, % | 16.6 | 1.2 | 0.5 |
| Have an outstanding loan, % | 72.6 | 55.9 | 68.3 |
| Social safety net program member/beneficiary<sup>3</sup>, % | 30.9 | 98.9 | 61.8 |
| National food support program beneficiary<sup>3</sup>, % | 15.9 | 96.8 | 34.9 |
| Pooled Wealth Quintile<sup>4</sup> |  |  |  |
| Q1, % | 23.8 | 17.9 | 15.1 |
| Q5, % | 15.9 | 13.8 | 34.7 |

<sup>1</sup>Names of the five districts and three countries from which data were derived are given in the column headings
<sup>2</sup>A household is considered to be food insecure if they fall under either category of mild/moderate/severe food insecurity.
<sup>3</sup>Social safety net programs include any income support programs such as the PMKisan Yojana in India. National food support programs include food distribution programs and school feeding programs.
<sup>4</sup>The numbers are the percentages of households in the lowest 20% (Q1) and highest 20% (Q5) of an asset-based wealth index generated on the data pooled across the five study districts

Many households did not have males present due to their migration for work. On average, female respondents were 37-42 years old with 3-5 years of education and males were 42-46 years old with 5-6 years of education. The female education disadvantage was largest in India (2.9 and 5.9 years for females and males respectively), whereas in Bangladesh females were slightly more educated than males (5.0 vs 4.6 years). Households in Bangladesh were also smaller and less likely to be female-headed compared to India or Nepal. In terms of other household characteristics, notable differences between countries included greater land ownership by females in Nepal, lower use of clean cooking fuel in Bangladesh, poor sanitation and relatively low agricultural and livestock or fish rearing involvement in India, and nearly full coverage of social safety net programs (both income and food) support programs in India, compared to much lower coverage, especially in Bangladesh. More than half of households were food insecure across study locations, with the highest prevalence in India (∼72%) and lowest in Nepal (∼55%).

### Diet quality metrics

The median duration of dietary recall using the GDQS application was 11 minutes in Banke, 12 minutes in Surkhet, 13 minutes in Nalanda, 18 minutes in Rajshahi, and 17 minutes in Rangpur. Total GDQS scores were very similar across countries and by gender, averaging 17-19 out of 49, which indicates low diet quality (**Figure 1, panels a-c**).

**Figure 1.**
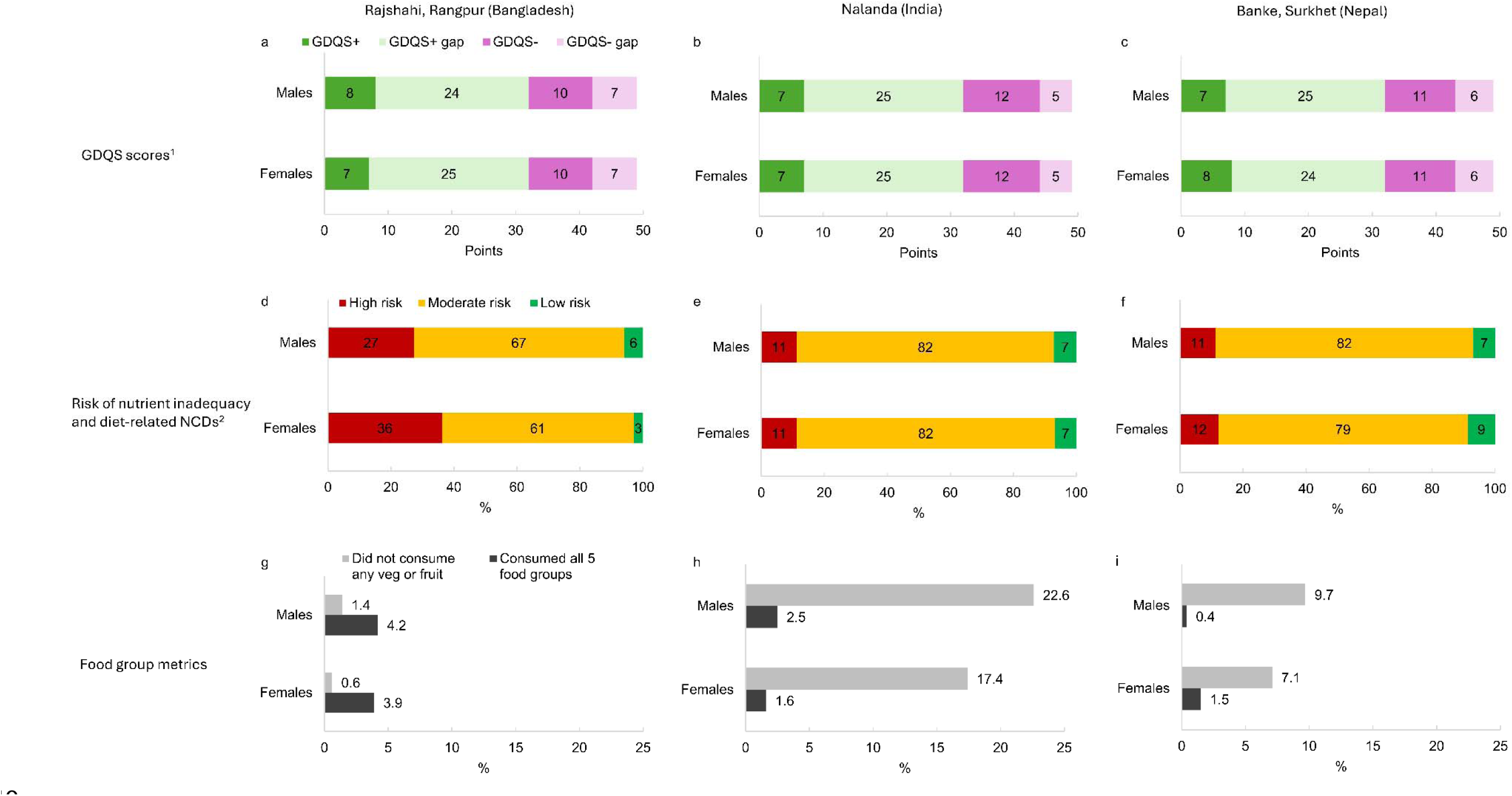
Diet quality metrics among male and female adults. ^1^The GDQS plus score ranges from 0-32 points and the GDQS minus score ranges from 0-17 points. The total GDQS score is the sum of the GDSQ+ and the GDQS-score. A higher score indicates better diet quality for all GDQS metrics. The GDQS+ and GDQS-gaps are the difference between the mean score and the maximum possible score (e.g. a GDQS+ score of 8 out of 32 has a GDQS+ gap of 24). ^2^The graphs (panels d, e, f) show the percentage of individuals in each risk category. Risk categories are derived from GDQS scores. A GDQS ≥23 is associated with a low risk of nutrient inadequacy and NCD-related outcomes, scores ≥ 15 and <23 indicate moderate risk, and scores <15 indicate high risk, based on the 24-hour reference period for which data were collected.

Categorizing the sample into groups of low, moderate, and high risk of nutrient inadequacy and diet-related NCDs, we found that only 3-9% of our samples were at low risk (Figure 1**, panels d-f**). The proportion of individuals at high risk for these poor health outcomes ranged from 11% to 36%, with males and females in Bangladesh having the highest risk, 27% and 36% respectively compared to 11-12% in India or Nepal. In terms of dietary diversity, however, nearly all individuals in Bangladesh consumed some vegetables or fruit whereas 1 in 5 adults in India did not. In both Nepal and India, the problem of zero vegetable or fruit intake was more common in males compared to females. Less than 5% of adults in all countries consumed all 5 food groups (at least one fruit, one vegetable, one pulse/nut/seed, one animal-source food, and one starchy staple) (Figure 1**, panels g-i**).

### Food group and food item intake

Among the 16 healthy GDQS food groups, citrus fruits and deep orange fruits, vegetables or tubers were rarely consumed across sites (Figure 2). Between 10% (India) and 20% (Bangladesh) of adults consumed other fruits and between 75% (India) and 100% (Bangladesh) consumed other vegetables. Legumes were consumed by about 40% (Bangladesh), 60% (India), and 80% (Nepal) of adults and were the main source of protein except in Bangladesh, where more than half of adults consumed fish. Whole grain consumption was much higher in India (∼100%) compared to Bangladesh (∼25%) or Nepal (∼20%). Poultry, dairy, egg, or meat intake was less common, though a quarter of adults in Bangladesh consumed eggs (vs. <10% in India and in Nepal) and 20% of adults in Nepal consumed poultry. There was very little intake of low-fat dairy in all three countries, but high-fat dairy was consumed in India (30%) and Nepal (20%), whereas in Bangladesh, intake was lower among females (∼10%) compared to males (20%). In terms of unhealthy foods, intake of refined grains and baked goods neared 100% in Bangladesh and Nepal, compared to 20% in India. Intake of red meat and processed meat in amounts considered unhealthy was very low in Bangladesh and Nepal and non-existent in India. Sweets were commonly consumed, especially by males in Bangladesh, but sugar sweetened beverages and juice were rarely consumed across study sites. Most individuals consumed white roots and tubers and around 10% consumed purchased fried foods.

**Figure 2.**
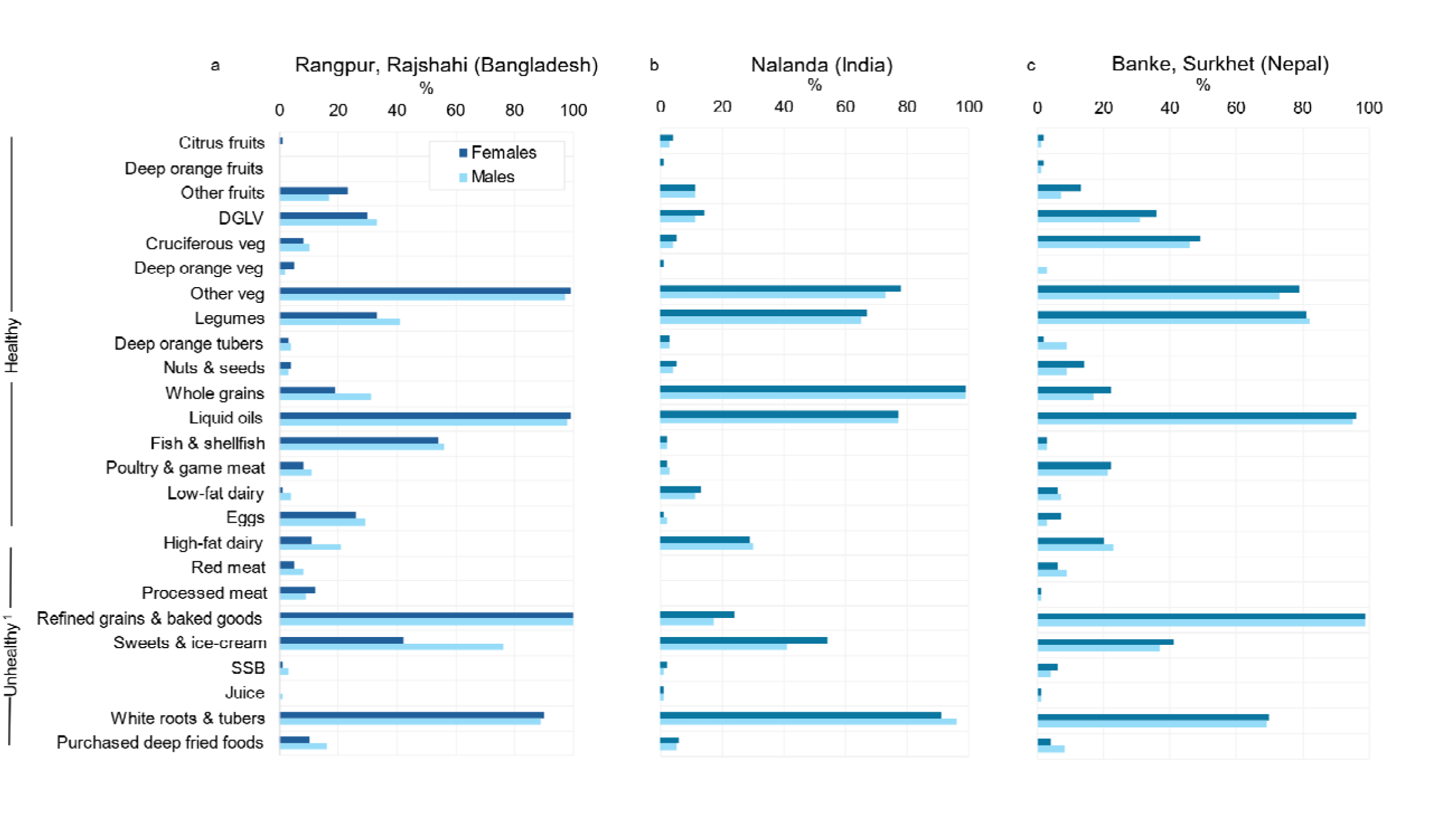
Percentage of male and female adults who consumed any amount of each GDQS food group in the last 24 hours. ^1^High-fat dairy and red meat are only considered unhealthy when consumed in high quantities.

Breaking down food groups into foods revealed that the most common vegetables consumed by adults aside from dark green leafy vegetables include onion, chili peppers, brinjal (eggplant), tomato, drumstick (moringa), ginger, and garlic (Figure 3**, panels a-c**). In India, brown rice and roti (a flatbread made from wheat flour) each made up about half of the whole grain category. In Nepal, there were multiple types of cruciferous and green leafy vegetables consumed. Among the GDQS unhealthy foods, potatoes were the predominant food consumed in the white roots and tubers category in all three countries (Figure 3**, panels d-f**). Refined grains included white rice and foods made from refined flours such as roti and instant noodles. The sweets category included sugar and biscuits.

**Figure 3.**
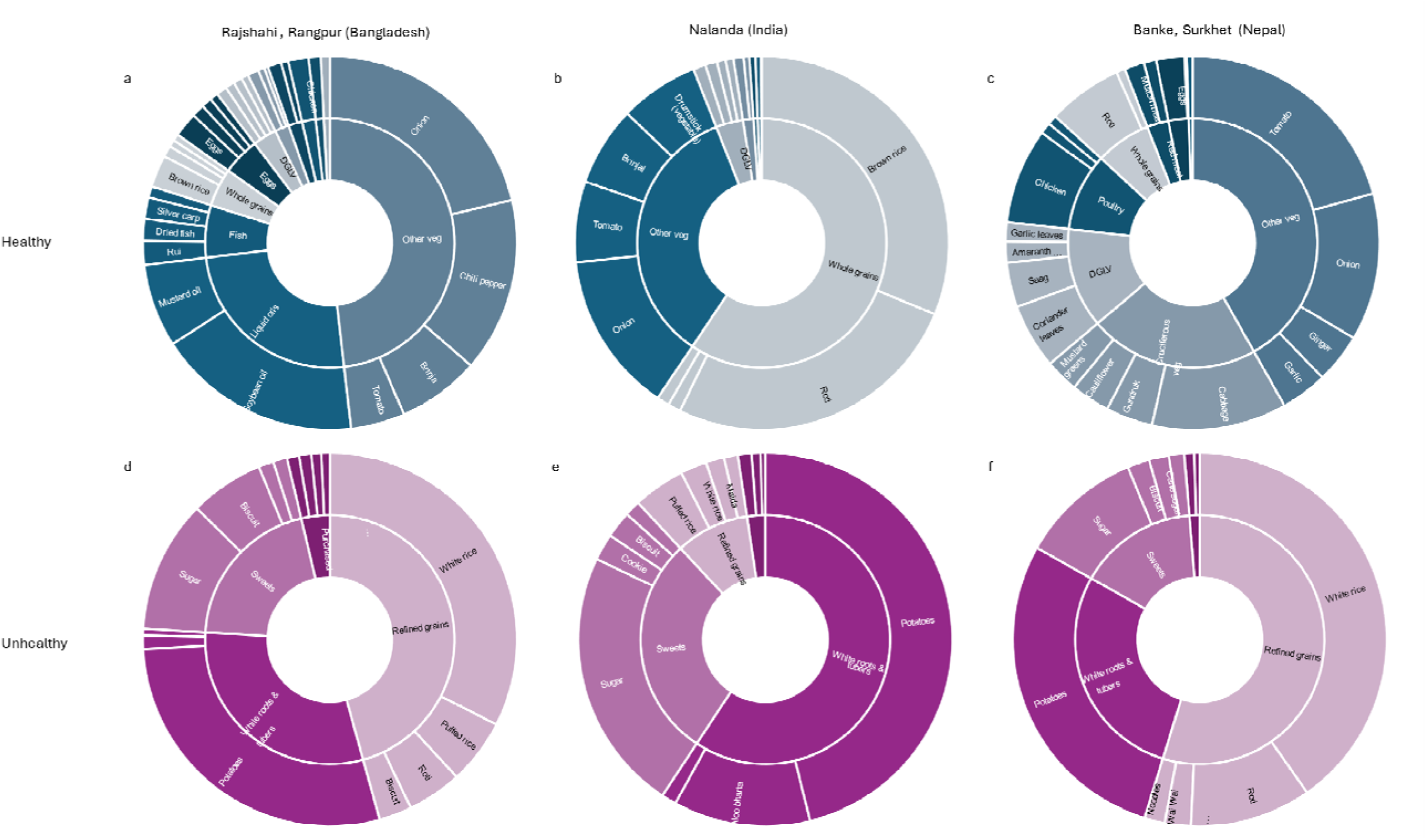
Healthy and unhealthy foods consumed for select food groups by adults. Data from males and females has been combined. The width of the outer blocks for each food or beverage is scaled according to the relative percent reporting consumption of that food. The size of each block in the inner circle reflects the sum of all percentages of the four mos frequently consumed foods for that food group. Text labels did not fit in all blocks, hence missing text where a low percentage of individuals consumed the food.

### Food intake across eating occasions

Eating patterns throughout the day differed by country and gender (Figure 4). Most adults consumed the three main meals of breakfast, lunch, and dinner, except in Nepal, where one-third of adults did not consume breakfast. Eating between the main meals or “snacking” was most common in Bangladesh, with around 40% of adults eating immediately after waking up before breakfast, 30% eating between breakfast and lunch, and 30% of females and 60% of males eating between lunch and dinner. Afternoon snacking was also common in Nepal, though higher in females compared to males.

**Figure 4.**
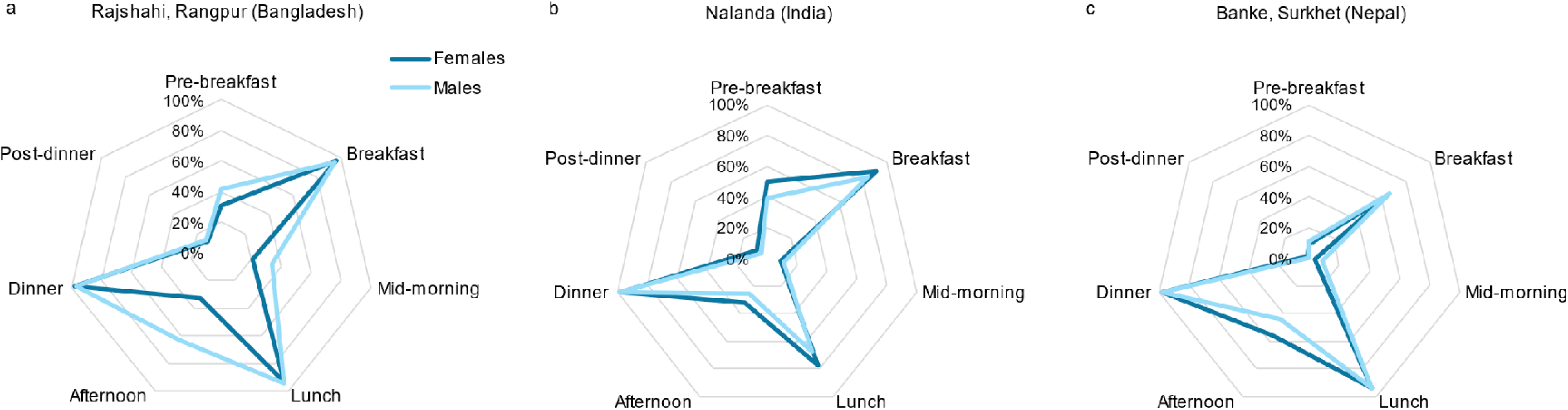
Percentage of male and female adults consuming any food at eating occasions throughout the day. Respondents were asked the following questions. Pre-breakfast: What was the first thing you ate/drank after you woke up yesterday (excluding water); Breakfast: What did you eat for breakfast?; Mid-morning: Did you eat anything between breakfast and lunch?; Lunch: What did you eat for lunch?; Afternoon: Did you eat anything between lunch and dinner?; Dinner: What did you eat for dinner?; Post-dinner: Did you eat anything after dinner

Looking at the foods and mixed food dishes consumed at each eating occasion, biscuits and tea were commonly consumed early in the day (Figure 5). The types of foods eaten at breakfast, lunch, and dinner were similar within countries. For example, in Bangladesh, bhaji (fried vegetables), vegetable curry, rice, and fish curry accounted for most of the main meals. In India, these staples included potato curry, vegetable curry, and rice (mostly for breakfast and lunch) or roti (mostly for dinner). In Nepal, where skipping breakfast was common, lunch and dinner staples included vegetable curry, rice, dal (lentil curry) and potato curry. This practice of a few staples being commonly consumed at main meals contrasts with a wider assortment of foods being consumed between main meals. For example, in Nepal, afternoon “snacks” included tea, chowmein, wai wai (instant noodles), chatpate (salty snacks), samosa/pakode (fried snacks), sometimes along with other foods consumed at main meals.

**Figure 5.**
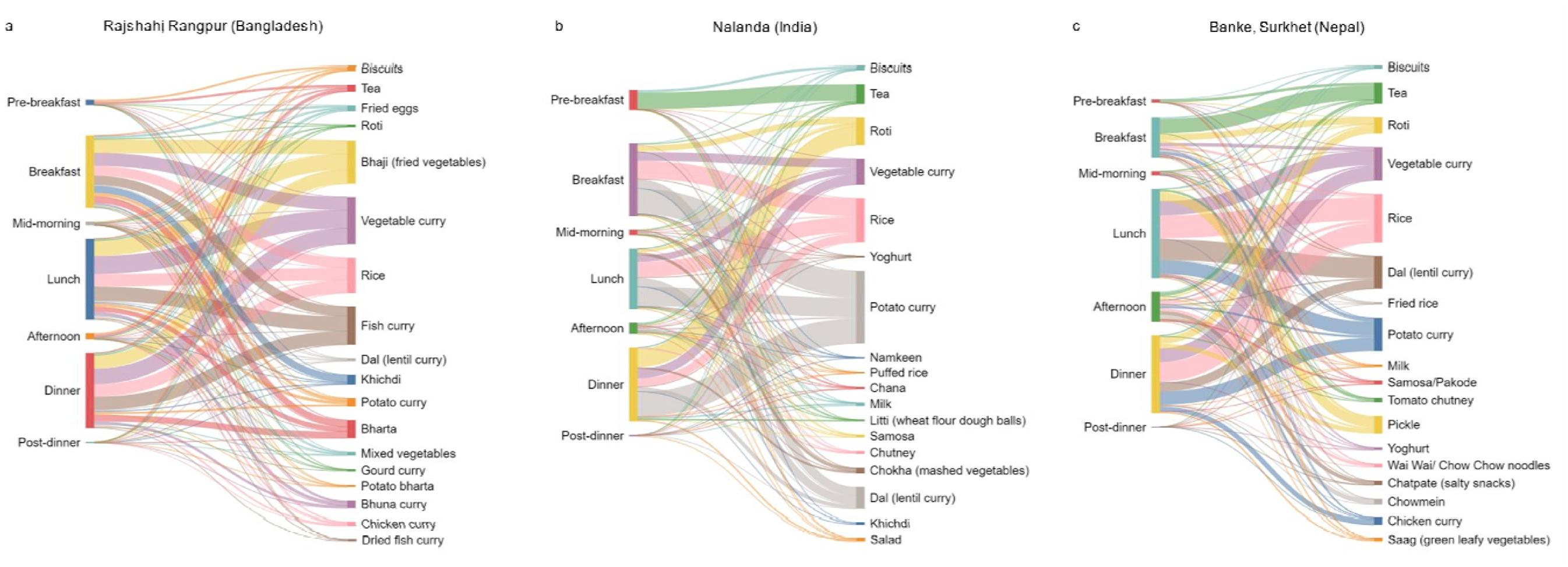
Food and mixed dishes consumed by adults at different eating occasions throughout the day. Foods and mixed dishes shown represent ∼80% of all the foods consumed in these locations. The foods/mixed dishes are scaled to the percentage of respondents consuming that food/dish for each eating occasion, e.g. in Nalanda (India) pre-breakfast consists of almost only tea, while breakfast consists of largely potato curry and rice, followed by Dal (lentil curry), vegetable curry, roti, and smaller amounts of other foods/mixed dishes.

## DISCUSSION

### Summary of main findings

We found that adult males and females in the surveyed districts in Bangladesh, India and Nepal had poor diet quality as indicated by low GDQS scores, which have been associated with moderate to high risk of nutrient inadequacy and diet-related NCDs. GDQS+ (healthy) scores were 7-8 out of a possible 32 points, and GDQS- (unhealthy) scores were 10-12 out of a possible 17 points, indicating a larger opportunity to improve overall diet quality through increased intake of healthy foods rather than to through decreased intake of unhealthy foods. To close the GDQS+ gap, adults could, in general, increase their intake of fruits, vegetables, and animal-source foods such as eggs, low-fat dairy, fish, or poultry, depending on which of these foods are culturally acceptable, available, accessible, and affordable. To close the GDQS-gap, adults could consume less refined grains and baked goods (e.g. reduce intake of white rice and replace with brown rice), white roots and tubers (e.g. potatoes), and sweets and ice cream (e.g. sugar and sweet biscuits).

In terms of eating patterns, we also found some differences between districts, with adults in the districts in Bangladesh, especially males, consuming more snacks between main meals than adults in India or Nepal. Finally, we found that the same few foods were commonly consumed at breakfast, lunch, and dinner, with 2-4 foods/dishes making up most of what was consumed at these main meals, whereas a greater variety of foods, often unhealthy, were consumed between main meals. Contrary to the general hypothesis that females have worse diets than males, we found few gender differences in dietary intake. Differences in diet patterns and meal timing across districts suggest that interventions that consider local norms are needed to improve diets.

### Comparison of GDQS findings with other studies

The GDQS metrics were validated using dietary data from surveys involving women of reproductive age in 10 African countries, and in China, India, Mexico, and the United States (13). The data from India were from the 2010-2012 rounds of the India Migration Study (IMS) in Telangana state and the 2009-2010 and 2010-2012 round of the Andhra Pradesh Children and Parents Study (APCAPS). Both IMS and APCAPS were from states in southern India, which are better off on most nutrition outcomes and determinants than Bihar (24–26), where our study was conducted. The mean GDQS, GDQS+, and GDQS-scores from the females in southern India were 23, 12, and 11 points respectively (27), compared to 19, 7, and 12 points respectively in our female sample in Nalanda. Poorer diets in northern India (Nalanda, Bihar) compared to southern India (Telangana and Andhra Pradesh) are due to relatively low intake of healthy foods in northern India. The Intake team at the Center for Dietary Assessment also computed GDQS scores from national rural representative data on adult diets collected by the Indian Council of Medical Research (ICMR) in 2009-2012 (28). The GDQS, GDQS+, and GDQS-scores from this data set were 18, 6, and 12, similar to the scores we found in Nalanda in 2023. We are not aware of studies from Bangladesh or Nepal reporting GDQS scores, though dietary surveys have been conducted in these countries and data from these surveys could be used to compute GDQS scores (10).

### Potential reasons for observed intake levels

We offer some reflections of why adults may be under-consuming healthy foods in rural South Asia, with additional findings from our agrifood systems survey results expected in forthcoming papers. Vegetarianism is common in the study population, especially in Nalanda (India sample). The GDQS only captures intake in the previous 24 hours, which may not pick up foods that are not consumed every day, however we also used a food frequency questionnaire to capture intake of 25 foods/food groups in the past week (not described in this paper) and found that 91% of adults (83% in Bangladesh, 99% in India, and 97% in Nepal) consumed no meat/fish/poultry/eggs in the past week. Cultural norms and preferences are critical determinants of food intake. A meal without rice would not be a meal in many households in South Asia. White rice (classified by GDQS into the unhealthy food group ‘refined grains’) is generally preferred over brown rice (classified into the healthy food group ‘whole grains’) for its appearance and taste, although most respondents in Nalanda reported consuming brown rice instead of white rice, which we cannot explain from our data but may simply be what is available to them. We also observed that most dairy consumed was high-fat dairy instead of low-fat dairy. This could reflect a preference for high-fat dairy, which is more satiating than low-fat dairy or low availability of low-fat dairy, which requires processing raw milk to remove a portion of the fat (29) and, related, consumption of unprocessed milk produced from owned livestock. Reassuringly, we did not observe high intake of sugar sweetened beverages and juices, which may not have yet penetrated rural villages in these study sites, although underreporting of unhealthy foods consumed is also a possibility. Our agrifood systems assessment included a market survey and captured information on food prices and perceptions about different foods and involvement of household members in different food-related tasks, thus further analysis of the drivers of food choice behaviors, including the role of food environments in shaping diets in this population, is planned.

### Heterogeneities in food intake behavior by location and gender

In terms of overall diet quality, we found few differences between males and females or across districts from three countries. However, despite mean GDQS scores being similar, there were three times as many individuals in Bangladesh at high risk of dietary inadequacy and diet-related NCDs compared to India or Nepal, and this was driven by relatively high intake of unhealthy foods in Bangladesh. A higher percentage of individuals in Bangladesh consumed processed meat, refined grains and baked goods, sweets and ice cream, and purchased deep fried foods compared to those in India or Nepal. Seemingly small differences in GDQS scores may mask larger differences in risk for poor health outcomes or in food intake. Based on this, we examined the breakdown of consumption level by food group, country, and gender (Supplementary Table 2). The main drivers of higher diet-related NCD risk, i.e. lower total GDQS points, in Bangladesh were high refined grain & baked goods consumption, low whole grain consumption, relatively low legume consumption, and relatively high sweets & ice cream consumption.

Our finding of minimal differences between males and females is supported by other recent work calling into question previous findings of pro-male intrahousehold food allocation South Asia (30). A study using data from rural Bangladesh collected in 2012 and 2016 reported that diets of men and women were equitable but suboptimal (31). After considering physiological differences in caloric needs and physical activity levels, the authors found that a pro-male allocation bias in energy intake disappears. Using GDQS scores, they also document that that both men and women had poor overall diet quality and that the difference between genders was minimal (women scored < 1% lower than men) and biologically insignificant. Similarly, an examination of Bangladesh Integrated Household Survey panel data by Ahmed et al. showed that the slightly better diets among men compared to women in 2011 were reversed by 2018, when women had more diverse diets than men (32). We did observe a few gender differences in types of foods consumed and in the timing of food intake. In Bangladesh, for example, intake of high fat dairy and sweets/ice cream as well as eating between main meals were much more common among males than females. In rural Bangladesh, women often do not have the freedom to move outside the home and thus do not have access to sweets shops and tea stalls (33).

### Strengths and limitations

We surveyed close to 6,000 adults from 4,000 households across five districts, and sampled households using a PPS approach so that our findings would be representative at the district level of rural households with an adolescent. The GDQS measurement approach allowed us to capture relatively detailed information on food intake within a broader food system survey where the GDQS was one of approximately twenty different modules capturing diets, food perceptions, food production, food and water insecurity, shopping habits, and intrahousehold task allocation among others. Our study is among the first to use the GDQS application and 3D-printed cubes to collect dietary data in a field-based setting (14), and fills an important data gap on dietary intake in South Asia. The main limitation of our survey is that it was conducted at a single timepoint and may not reflect variations in dietary intake throughout the year.

### Policy implications and research needs

Urgent actions are required to address the poor quality of diets in rural South Asia found in this study. While this paper describes what adults eat and when they eat certain foods throughout the day, a deeper understanding of drivers of food choices in each context is needed to design effective interventions to increase intake of healthy foods and limit intake of unhealthy foods. Nonetheless, policy insights can be drawn from the current findings. First, recognizing that changing behavior is challenging, it is critical to understand typical eating behaviors and design interventions that are acceptable and nudge consumers toward better habits rather than recommending total elimination of unhealthy foods or introducing unfamiliar foods. For example, we found that within snacking occasions, a variety of healthy and unhealthy foods were consumed; substituting unhealthy snacks with healthy snacks that are already available and consumed may be a more effective approach than recommending healthy foods that are not normally consumed. Second, we found that around 1 in 3 adults in Nepal did not consume any food before lunch and, for those who did consume breakfast, it was mostly tea and light snacks. Studies have reported that skipping breakfast is associated with lower cognitive function and work efficiency (34) as well as higher risk of obesity and diabetes (35,36). Investments in understanding morning eating practices in Nepal might help to assess whether specific benefits in this population may result from an intervention to increase food consumption early in the day.

## Conclusion

This study adds to the limited evidence on food intake and intake patterns in rural South Asia. The type of data we collected could be a useful source of information for updating food based dietary guidelines and shaping recommendations to consider cultural factors and preferences. This would require gathering similar data in all key regions of a given country to identify critical dietary gaps (e.g. low intake of fruit and vegetables and low-fat dairy; over-reliance on refined cereals; role of snacks and breakfast) and provide guidance to design regional and local solutions to address them. Investments in collection of quantitative dietary data representative at sub-national levels at different times of year, alongside collection of data on factors that shape food choices are essential in South Asia. Together with research studies on tailored solutions to tackle specific drivers of food choice, these can provide insights on how to shift dietary patterns to achieve culturally acceptable sustainable healthy diets for all.

## Data Availability

All data produced are available on the Harvard Dataverse, with links to the datasets available at: https://www.cgiar.org/news-events/news/open-access-agrifood-system-data-from-4000-households-across-bangladesh-india-and-nepal/

---

**Supplementary Table 1.** GDQS Food Groups and Scoring

| Scoring Classification | Food Group | Categories of Consumed: Amounts (g/day) |  |  |  | Points Assigned |  |  |  |
| --- | --- | --- | --- | --- | --- | --- | --- | --- | --- |
|  |  | Low | Middle | High | Very High | Low | Middle | High | Very High |
| Healthy | Citrus fruits | <24 | 24–69 | >69 |  | 0 | 1 | 2 |  |
|  | Deep orange fruits | <25 | 25–123 | >123 |  | 0 | 1 | 2 |  |
|  | Other fruits | <27 | 27–107 | >107 |  | 0 | 1 | 2 |  |
|  | Dark green leafy vegetables | <13 | 13–37 | >37 |  | 0 | 2 | 4 |  |
|  | Cruciferous vegetables | <13 | 13–36 | >36 |  | 0 | 0.25 | 0.5 |  |
|  | Deep orange vegetables | <9 | 9–45 | >45 |  | 0 | 0.25 | 0.5 |  |
|  | Other vegetables | <23 | 23–114 | >114 |  | 0 | 0.25 | 0.5 |  |
|  | Legumes | <9 | 9–42 | >42 |  | 0 | 2 | 4 |  |
|  | Deep orange tubers | <12 | 12–63 | >63 |  | 0 | 0.25 | 0.5 |  |
|  | Nuts and seeds | <7 | 7–13 | >13 |  | 0 | 2 | 4 |  |
|  | Whole grains | <8 | 8–13 | >13 |  | 0 | 1 | 2 |  |
|  | Liquid oils | <2 | 2–7.5 | >7.5 |  | 0 | 1 | 2 |  |
|  | Fish and shellfish | <14 | 14–71 | >71 |  | 0 | 1 | 2 |  |
|  | Poultry and game meat | <16 | 16–44 | >44 |  | 0 | 1 | 2 |  |
|  | Low-fat dairy | <33 | 33–132 | >132 |  | 0 | 1 | 2 |  |
|  | Eggs | <6 | 6–32 | >32 |  | 0 | 1 | 2 |  |
| Unhealthy in excessive amounts | High fat dairy<sup>1</sup> (in milk equivalents) | <35 | 35–142 | >142–734 | >734 | 0 | 1 | 2 | 0 |
|  | Red meat | <9 | 9–46 | >46 |  | 0 | 1 | 0 |  |
| Unhealthy | Processed meat | <9 | 9–30 | >30 |  | 2 | 1 | 0 |  |
|  | Refined grains and baked goods | <7 | 7–33 | >33 |  | 2 | 1 | 0 |  |
|  | Sweets and ice cream | <13 | 13–37 | >37 |  | 2 | 1 | 0 |  |
|  | Sugar-sweetened beverages | <57 | 57–180 | >180 |  | 2 | 1 | 0 |  |
|  | Juice | <36 | 36–144 | >144 |  | 2 | 1 | 0 |  |
|  | White roots and tubers | <27 | 27–107 | >107 |  | 2 | 1 | 0 |  |
|  | Purchased deep-fried foods | <9 | 9–45 | >45 |  | 2 | 1 | 0 |  |

<sup>1</sup> Hard cheese should be converted to milk equivalents using a conversion factor of 6.1 when calculating total consumption of high-fat dairy for the purpose of assigning a GDQS consumption category.

**Supplementary Table 2.**
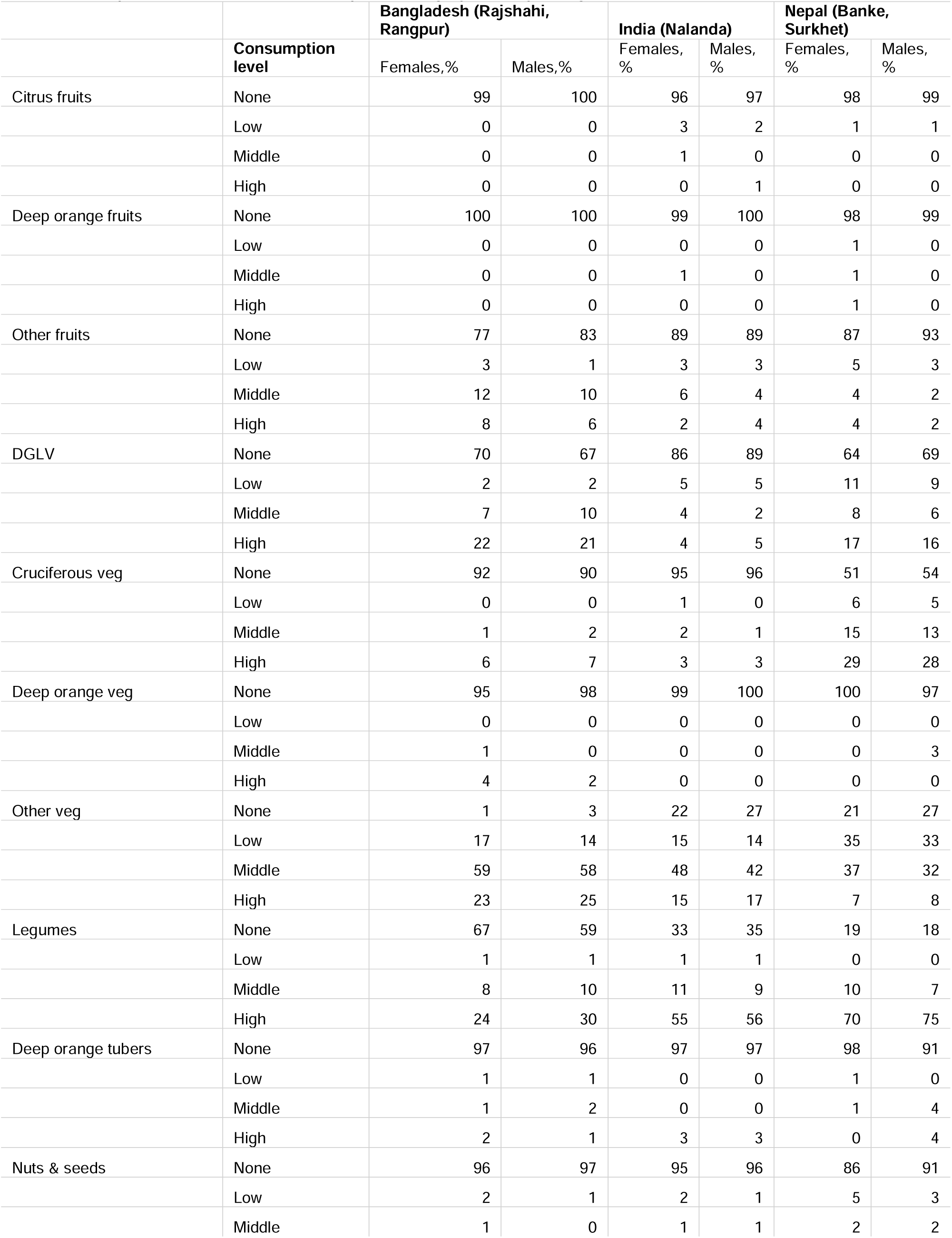
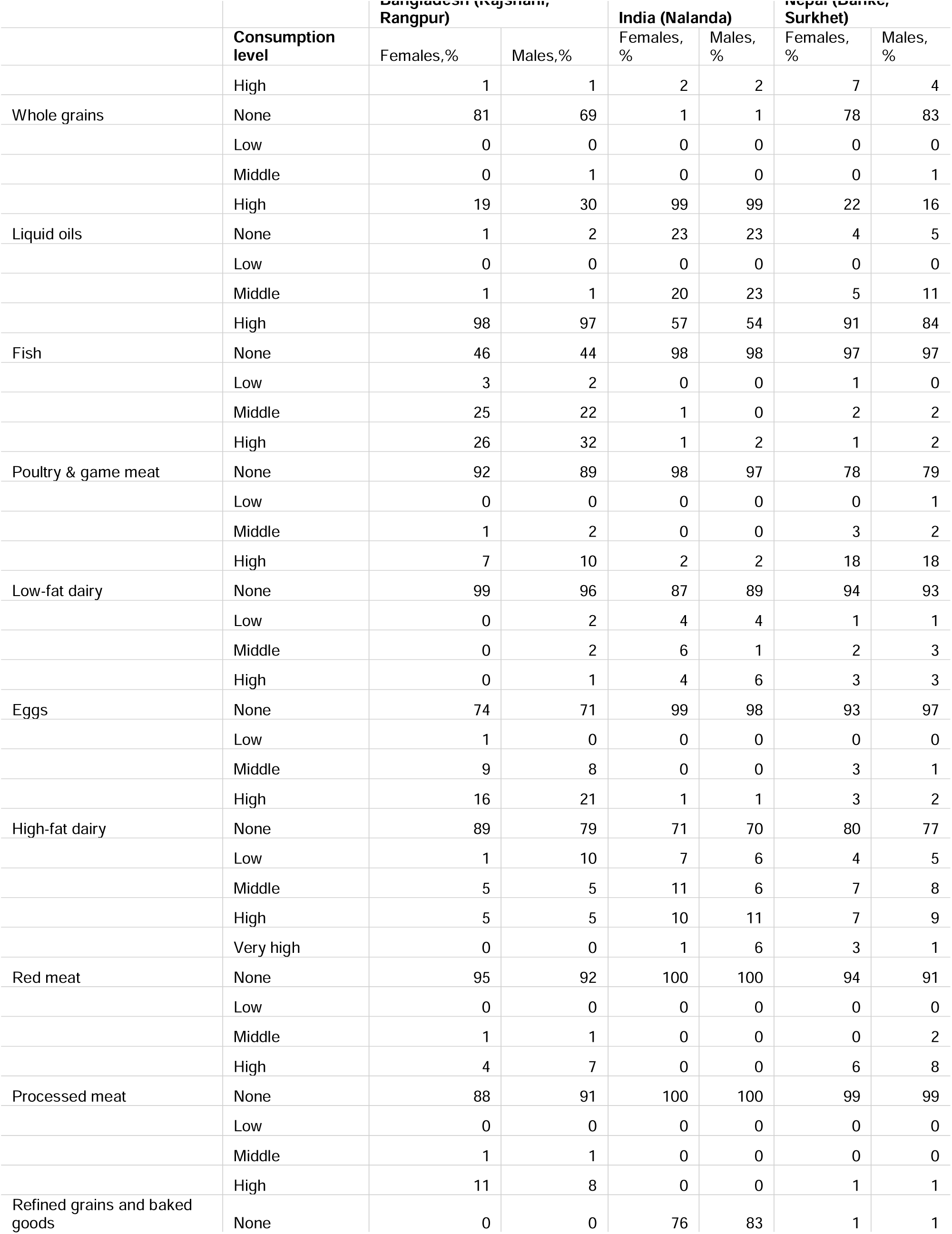
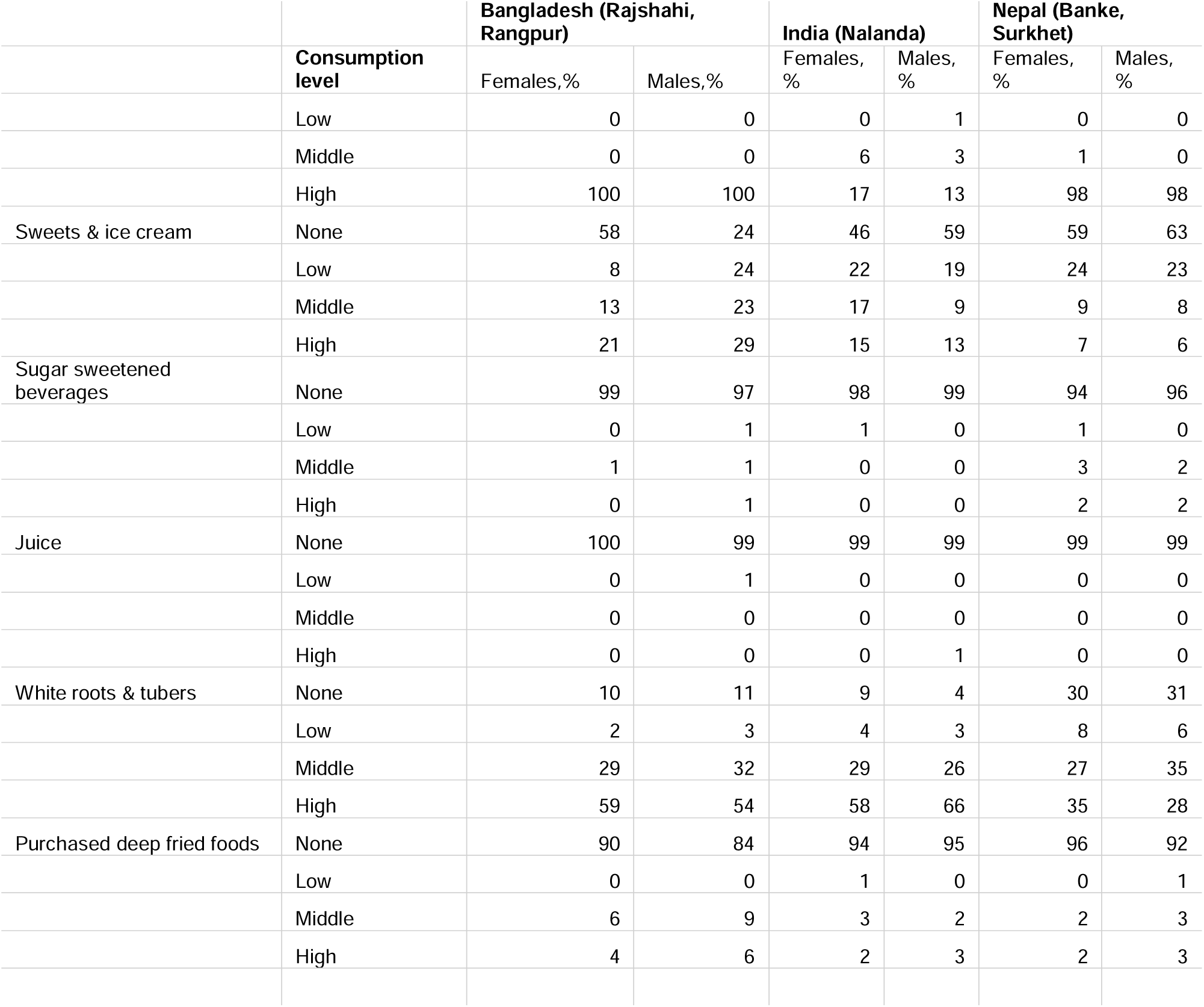
Consumption level by food category, country, and gender

## Notes

### Competing Interest Statement

The authors have declared no competing interest.

### Funding Statement

We acknowledge all funders who supported this research through their contributions to the CGIAR Trust Fund: www.cgiar.org/funders/.

